# Incorporating efficacy data from initial trials into subsequent evaluations: Application to vaccines against respiratory syncytial virus

**DOI:** 10.1101/2023.03.27.23287639

**Authors:** Joshua L. Warren, Maria Sundaram, Virginia E. Pitzer, Saad B. Omer, Daniel M. Weinberger

## Abstract

**Background:** When a randomized controlled trial fails to demonstrate statistically significant efficacy against the primary endpoint, a potentially costly new trial would need to be conducted to receive licensure. Incorporating data from previous trials might allow for the conduct of more efficient follow-up trials to demonstrate efficacy, speeding the availability of effective vaccines.

**Methods:** Based on the outcomes from a failed trial of a maternal vaccine against respiratory syncytial virus (RSV), we simulated data for a new Bayesian group-sequential trial. The data were analyzed either ignoring data from the previous trial (i.e., weakly informative prior distributions) or using prior distributions that incorporate the historical data into the analysis. We evaluated scenarios where the efficacy in the new trial was the same, greater than, or less than the efficacy in the original trial. For each scenario, we evaluated the statistical power and type I error rate for estimating the vaccine effect following interim analyses.

**Results:** If a stringent threshold is used to control the type I error rate, the analyses that incorporated historical data had a small advantage over trials that did not. If control of type I error is less important (e.g., in a post-licensure evaluation), the incorporation of historical data can provide a substantial boost in efficiency.

**Conclusions:** Due to the need to control the type I error rate in trials used to license a vaccine, the incorporation of historical data provides little additional benefit in terms of stopping the trial early. However, these statistical approaches could be promising in evaluations that use real-world evidence following licensure.

## INTRODUCTION

When testing new vaccines, there is a need to ensure that the product is safe and effective. This is typically accomplished through a randomized controlled trial where participants are randomly assigned to receive the new vaccine or a placebo [1]. These trials have become increasingly large and costly [2] and have stringent criteria for success. This can lead to delays in introducing potentially lifesaving vaccines or even forestall the development of promising vaccines that fail to meet strict guidelines for approval. An example of this comes from a maternal vaccine against respiratory syncytial virus (RSV) that was tested in a phase 3 clinical trial [3]. RSV is a leading cause of hospitalization in infants globally [4]. The vaccine failed to show efficacy against the primary clinical outcome of medically significant RSV (efficacy of 39.4%, with a 97.52% confidence interval (CI) that ranged from −1.0% to 63.7%). However, examining several of the secondary clinical outcomes, there was clear evidence of a beneficial effect against RSV hospitalizations. Nevertheless, because the CI for the primary outcome included zero, the trial was deemed a failure, and licensure of the vaccine and further testing were not pursued. To be licensed, this vaccine would likely need to again undergo testing in a large phase 3 clinical trial.

Even if a vaccine is successfully licensed based on data from a phase 3 randomized trial, there is often a requirement from regulators that manufacturers monitor safety and effectiveness in real-world studies [5]. In such studies, the goal is to estimate the effects of the vaccine and detect safety signals in a larger, less restricted population.

Both randomized controlled trials and post-licensure evaluations are typically conducted as stand-alone analyses that do not incorporate knowledge obtained from prior trials or evaluations. In the case of a failed phase 3 trial in which there is evidence of a beneficial but not statistically significant effect of vaccination, one possibility to speed licensure of the vaccine would be to incorporate data from the initial trial into the analysis of data from a new, smaller trial [6,7]. This could be conducted as a traditional trial with a fixed sample size or, potentially more efficiently, as a group sequential Bayesian trial [8]. With group sequential trials, vaccine efficacy (VE) is evaluated on an ongoing basis during interim evaluations, and the trial is stopped when sufficient evidence of efficacy has been collected [8,9]. In the case of a post-licensure evaluation of vaccine effectiveness, estimates from the phase 3 trial could be integrated into analyses that confirm the study results of safety or efficacy. Relatedly, the trial results could be integrated into analyses of endpoints that are different from those evaluated in the trial (e.g., rare but severe outcomes like intensive care unit admission or death) or risk groups for which it was not possible to obtain robust estimates in the trial. For example, a phase 3 trial of an RSV vaccine in older adults included a subgroup analysis of efficacy in adults over the age of 80 years [10,11]. There was a high degree of uncertainty in these estimates, which partially led to a weaker recommendation for the vaccine from the US Advisory Committee on Immunization Practices. Incorporating real-world evidence with the trial data would allow for more rapid and efficient evaluations of these crucial endpoints of interest.

There is precedent for using group sequential designs and Bayesian analyses in vaccine trials, particularly in the context of public health emergencies, such as for trials of vaccines against SARS-CoV-2 and Ebola. A group sequential Bayesian design was used for the phase 3 portion of the trial that led to licensure of the BNT162b2 vaccine (Pfizer-BioNTech) against SARS-CoV-2 [12]. In the study protocol, the investigators provided detailed justification of the criteria for success and futility based on a variety of expected levels of vaccine efficacy and sample sizes. Similarly, a trial conducted of the Ad26.COV2.S vaccine (Janssen) against SARS-CoV-2 used a frequentist group sequential design [13]. The original trial of the Novavax maternal RSV vaccine also had originally planned to use a group sequential design, which was abandoned following the interim analyses [16].

Using novel designs of clinical trials presents certain challenges, particularly when working in a Bayesian analysis framework. Critical study characteristics, such as power, type I error, and expected sample size can be influenced by the selected prior distribution(s) for the vaccine efficacy. The incorporation of secondary data, as is proposed in this study, further complicates the planning of these trials. Therefore, the use of data simulations performed during the study design phase are critical to demonstrate the likely characteristics of the study and the influence of prior distributions on the statistical inference [14–16].

In this paper, we focused on the application of methods that incorporate data from previous trials into the design of a hypothetical follow-up to a failed phase 3 trial of a maternal vaccine against RSV [3]. Using simulation analyses, we explored the advantages and potential pitfalls of incorporating estimates of VE from a previous trial into the analysis of data from a new trial.

## METHODS

### Overview of analyses and definitions

The goal of these analyses was to evaluate the performance of different approaches for analyzing data from a hypothetical new trial of the Novavax maternal vaccine against RSV. We used data from a completed phase 3 trial of the Novavax maternal RSV vaccine (“Prepare trial”) as the historical data to inform the prior distributions for some of our analyses [3]. The original trial did not demonstrate statistically significant efficacy against the primary clinical endpoint, but there was efficacy against secondary endpoints. We focused on a secondary endpoint in the trial that showed particular promise and that could be considered as a primary endpoint in the hypothetical subsequent trial seeking licensure: VE against hospitalized RSV lower respiratory tract infections (RSV LRTI) [3]. We simulated data for a hypothetical new trial evaluating the risk of hospitalization for RSV LRTI in infants of vaccinated mothers compared to infants of mothers who received the placebo. Mirroring the original trial, the main estimand of interest was a risk ratio, defined as the proportion of infants hospitalized for RSV LRTI in the vaccinated group divided by the proportion of infants hospitalized for RSV LRTI in the placebo group.

Our simulated trial used a Bayesian group-sequential design. We simulated data mirroring the original trial [3], in which the vaccine was given to pregnant women between 27 and 37 weeks of gestational age (mean 32 weeks), and the primary endpoint was the number of cases of RSV LRTI in the first 90 days of life; therefore, there were ∼5 months between vaccine receipt and assessment of the primary endpoint. We considered a trial that enrolled in the Northern and Southern Hemispheres prior to the respective RSV seasons, with a maximum of 6000 total participants enrolled over three years, i.e. up to six groups of 1000 participants each would be enrolled every six months. Interim analyses were planned after each 1000 participants, which would allow for the trial to be halted before the other Hemisphere began enrollment for the season. We assumed participants would be randomized 1:1 to receive a vaccine or placebo. Several different prior distributions for the effect of vaccination were evaluated (detailed below), including prior distributions that incorporated data from the previous (failed) trial and minimally informative prior distributions.

The simulated trials were stopped early if the posterior probability of the VE being > 0 was greater than a pre-specified threshold at each interim evaluation (i.e., P(VE > 0 | data) > (1 - α_interim_)). As described below, we evaluated different values of α_interim_ to control the overall type I error in the sequential trials. The same stopping criterion, α_interim_, was used at all interim analyses and in the final analysis. There was no stopping rule based on futility or safety, as would be the case in a real trial.

### Data from the previous trial

In the Prepare trial, VE against hospitalized RSV LRTI, measured at day 90 after birth in a per protocol analysis, was 44.4% (95% CI: 19.6%, 61.5%) (second table in the original paper [9]). This estimate of VE was based on 53 cases out of 1430 participants in the placebo group and 57 cases out of 2765 participants in the vaccinated group.

### Generation of simulated data

The primary outcome for each individual was a binary variable describing whether or not they were hospitalized with RSV LRTI during the 90-day follow-up period. The binary outcome data from a single simulated trial for person *j* in treatment arm *v* (i.e., Case_status*_vj_*) were generated as

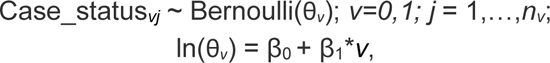

where *v=0* corresponds to the placebo group and *v=1* to the vaccinated group; *n=n_0_* + *n_1_* is the total number of individuals enrolled in the trial; θ*_v_* is the true probability of hospitalization for RSV LRTI for every individual in treatment arm *v*; exp(β_1_) represents the risk ratio of hospitalization for RSV LRTI in the vaccinated group vs placebo group; and 1-exp(β_1_) is the VE, which we report as a percentage. When generating the data, β_0_ is fixed at ln(0.037) based on the proportion of infants in the placebo arm of the Prepare trial who were hospitalized with RSV LRTI (i.e., 0.037, or 3.7%), and β_1_ is assigned a fixed value as described for the three scenarios below.

We simulated data representing three scenarios by changing the value of β_1_ from the data-generating model:

1. There is no effect of the vaccine in the new trial (VE = 0%; β_1_ = 0).
2. The VE in the new trial is the same as in the Prepare trial (VE=44.4%; β_1_ = ln(0.556)).
3. The VE in the new trial is higher than in the original trial (VE=90%; β_1_ = ln(0.100)).

For each scenario, we generated 500 simulated datasets, with up to 6000 participants.

### Analysis

All analyses of the simulated data were carried out in the Bayesian setting. The observed number of cases among all individuals in treatment arm *v* was modeled using Poisson regression such that

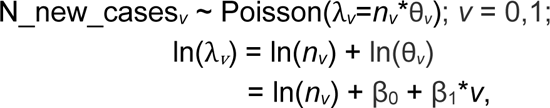

where N_new_cases*_v_* was the total number of RSV LRTI hospitalizations observed in treatment arm *v* from the new/simulated trial; λ*_v_* was the expected number of cases; and all other terms have been previously described. A Poisson regression model was used to mirror the analysis in the original Prepare trial.

For analyses that incorporate historical data, the VE from the original trial was also re-estimated using Poisson regression such that

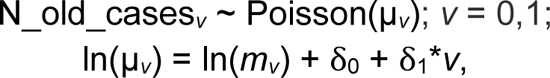

where N_old_cases*_v_* is the total number of RSV LRTI hospitalizations observed in treatment arm *v* from the original Prepare trial; µ*_v_* is the expected number of cases; *m_v_* is the number of individuals in treatment arm *v* from the original Prepare trial; and δ_0_, δ_1_ represent the risk in the placebo group and the effect of vaccination, respectively, based on data from the original trial. The β_0_, β_1,_ δ_0_, and δ_1_ parameters were all estimated simultaneously when incorporating historical data into the analysis.

We evaluated the performance of several different prior distributions for β_0_ and β_1_ (**Table S1**). The prior distributions we tested were:

a. *Weakly informative prior distributions, no historical data used.* This scenario most closely resembles a typical clinical trial, where there is no prior information incorporated into the new analysis. The prior distributions in this scenario are β_0,_ β_1_ ∼ Normal(0, 100^2^).
b. *Skeptical prior for* β_1_*, no historical data used.* This specification is similar to (a), but with a more informative prior distribution selected to penalize extreme VE values (i.e., large beneficial or adverse effects of the vaccine [17]). Specifically, the prior distribution is given as β_1_ ∼ Normal(0, 3.32) which corresponds to a <5% prior probability that the risk ratio is less than 0.05 or greater than 20 (i.e., a risk ratio roughly between −3 and 3 on the log scale). For the intercept, we still use a weakly informative prior distribution: β_0_ ∼ Normal(0, 100^2^).
c. *Commensurate prior for* β_1_ *using historical data*. For this method, originally developed by Hobbs [6], the prior distributions for the current trial parameters are centered at the corresponding parameters from the historical trial such that β_0_ ∼ Normal(δ_0_, σ^2^_0_) and β_1_ ∼

Normal(δ_1_, σ^2^), with weakly informative priors specified for the historical data parameters (i.e., δ_0_, δ_1_ ∼ Normal(0, 100^2^)). Estimation of the variance parameters (i.e., σ^2^, σ^2^) is driven by comparing the level of agreement between the historical and current datasets with respect to these parameters. A large variance suggests that only data from the current trial are used to estimate the corresponding parameter, while a small variance results in more information-borrowing from the historical data.

We evaluated several prior distributions for these variance parameters, as they control the level of information-sharing between the historical and current trials. Specifically, we tested σ^2^, σ^2^ ∼ Inverse-Gamma(0.01,0.01), Inverse-Gamma(1,1), and Inverse-Gamma(3,2). Alternatively, we specified the prior distribution on the standard deviation scale for these parameters such that σ_0_, σ_1_ ∼ Uniform(0,2), Uniform(0,100), and half-Cauchy [18]. We quantified the amount of information-sharing between the historical and current trials as a transformation of the variance parameter from the commensurate prior (exp(-σ^2^1)). If the variance was small, that indicated increased sharing, and the transformed value was close to 1. If the variance was large, this indicated decreased sharing, and the value approached 0.

All models were fit with the rjags package in R, where we collected 10,000 samples from the joint posterior distribution after discarding the first 10,000 prior to convergence of the model.

### Operating characteristics of the trial (frequentist power)

For all analyses, the goal was to estimate the true VE. Because we worked in the Bayesian setting, we used posterior tail probabilities to describe statistical significance. The main quantity of interest was the posterior probability that VE > 0%. We also considered a more stringent endpoint with the posterior probability that VE > 30%. Across the 500 simulated trials, statistical power was estimated as the *proportion of trials* that had an interim or final analysis where the posterior probability was greater than (1 – α_total_). Without adjustment for type I error, α_interim_ was set to 0.05 (i.e., one-sided test). To control the overall type I error for the sequential trials (α_total_≤0.05), we calibrated the value of α_interim_ in simulated trials where the true VE was 0% or 30%, depending on the specified endpoint. A value of α_interim_ was selected by varying α_interim_ until approximately 5% of simulated trials were declared a success by the time the maximum population size (6000 participants) was reached. The same α_interim_ (e.g., α_interim_=0.015) was used at each interim analysis and the final analysis.

### Availability of code

All analyses were completed using R v4.1.2, using the rjags package for analysis [19,20]. All code required to replicate these analyses is available at https://github.com/weinbergerlab/sequential_bayesian_trials.

## RESULTS

As data accrued in a group sequential trial, the estimate of efficacy was updated, as shown in an example simulated trial where the VE was evaluated after every 200^th^ participant completed the trial (**Figure 1**). With smaller samples sizes, the prior distribution (**Table S2**) and use of historical information had a larger influence on the inference, resulting in narrower credible intervals when the true VE was 0% or 44.4% (**Figure 1**). When the true VE was 90% (greater VE than in the historical data), the incorporation of historical information resulted in wider credible intervals at low sample sizes. When there were more than 2000-3000 participants across both treatment arms (∼50 cases, **Figure S1**), the estimates were similar regardless of the choice of prior distribution or the use of historical information (**Figure 1**). When the observed data aligned more closely with the data from the original trial, more sharing of information occurred, as expected (**Figure S2**).

**Figure 1.**
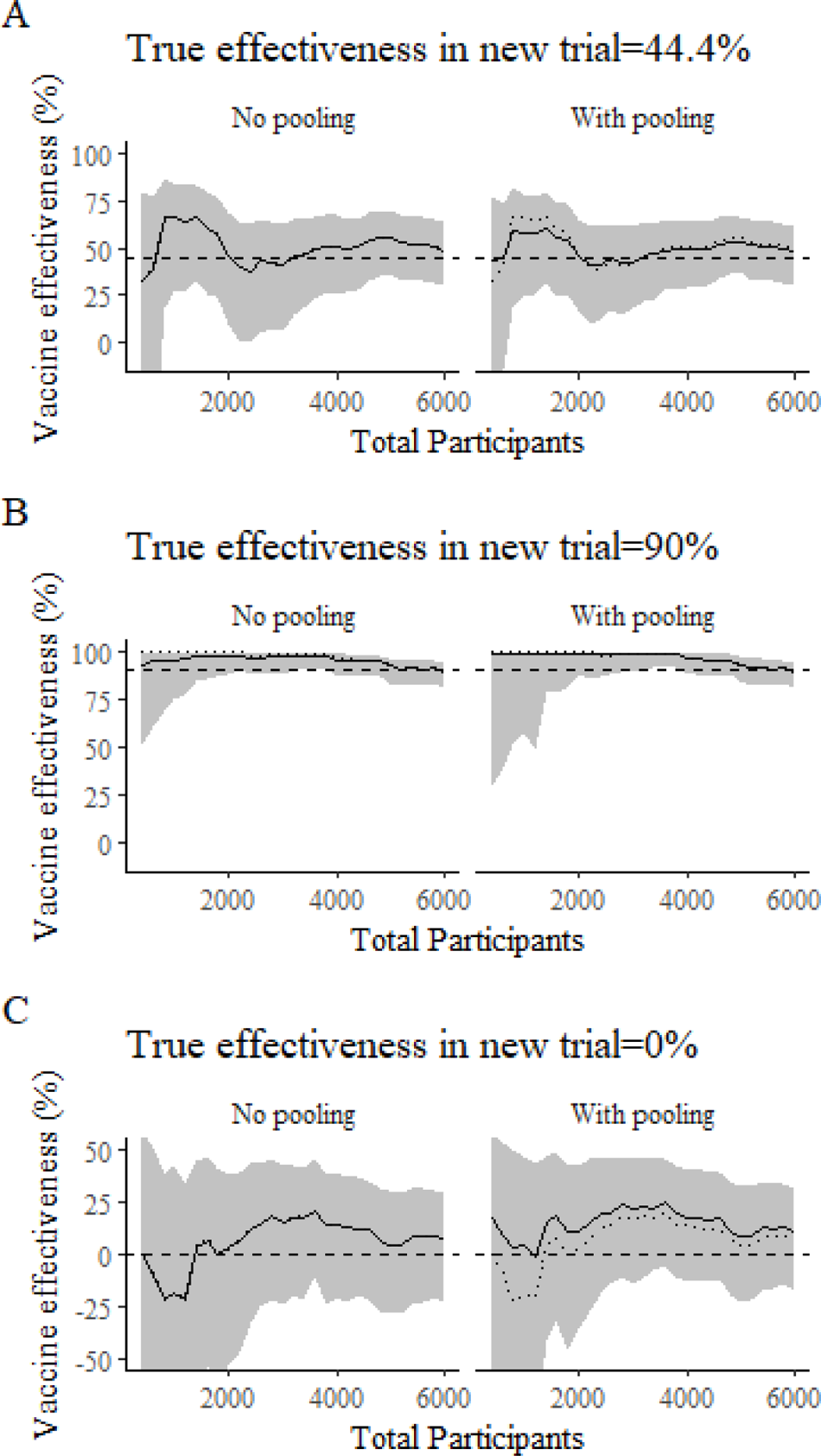
Evolution of the estimated risk ratio in a single trial as more participants complete the trial. The panels on the left are calculated without using historical data and a skeptical prior on the vaccine effect (i.e. β_1_ ∼ N(0,3.32)). The panels on the right represent estimates from a model that uses a commensurate prior to incorporate estimates from a previous trial; the prior on the vaccine effect is Normal(δ_1_,σ^2^); σ^2^∼InverseGamma(0.01,0.01). In this example, the VE is calculated on a continuous basis after every 200^th^ participant completes the study, starting with the 400^th^ participant. The posterior median estimated values for the risk ratio (1 – VE) are shown with the black line, the 95% credible intervals are represented by the gray shaded area. The dotted line shows the risk ratio calculated empirically based on the observed data only. The dashed line indicates the true VE for each panel (44.4%, 90%, 0%). In Panel A, the true VE is 44.4% (concordant with the historical estimate), in panel B, the true VE is 90% (greater than the historical estimate), and in panel C, the true VE is 0% (less than the historical estimate). Note the y-axis for panel C is different than the y-axes for panels A and B.

Examining the expected trial performance across 500 simulated trials, the analyses that incorporated historical data performed only slightly better in terms of statistical power (i.e., the proportion of simulated trials where a significant effect was correctly identified; **Figure 2, Figure S3**) compared with methods that did not use historical data. This is because of the need to use a more stringent threshold for α_interim_ to reduce the type I error rate when incorporating historical data (**Table S2**).

**Figure 2.**
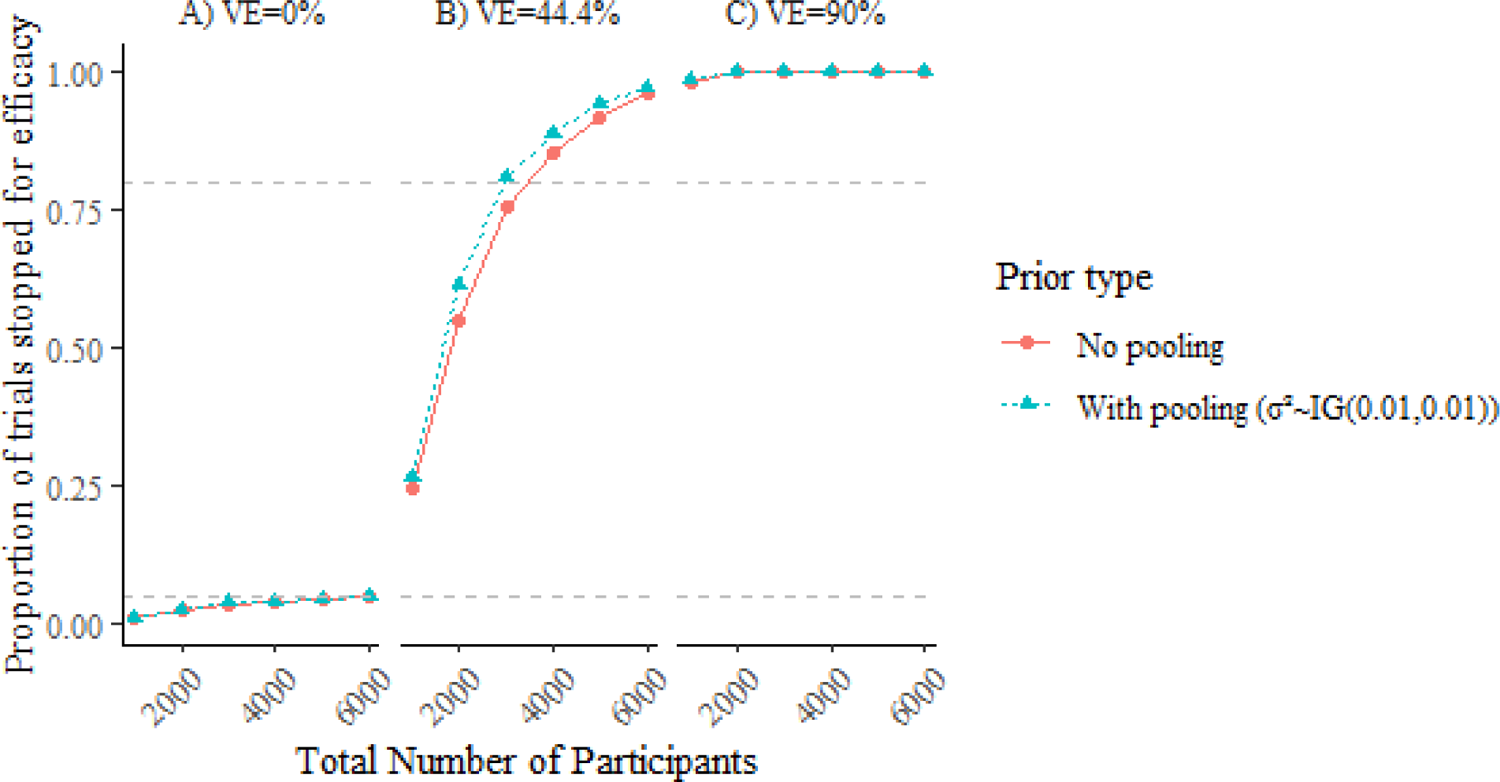
Cumulative proportion of 500 simulated trials that are declared a success (“power”) using a threshold for stopping (α_interim_) that is adjusted to maintain a type I error rate of 5% for the study (α_T_). Success is defined based on a posterior probability that VE > 0% is greater than or equal to 1-α_interim_. VE is evaluated after every 1000 participants complete the follow-up period. From left to right, we consider scenarios with a “true” vaccine efficacy of 0%, 44.4%, and 90%. The horizontal dashed lines are at 0.05 and 0.80, which represent arbitrary thresholds for acceptable type I error rates and power, respectively. The value of α_interim_ is adjusted so that the proportion of trials stopped for efficacy at or before 6000 participants is 0.05 when the true VE=0% (left panel). Two analyses are shown: one in which only data from the new trial is used (No pooling; red dots) and one where a commensurate prior is used to pool the data from the simulated new trial and the original trial (With pooling; blue triangles).

Trials tended to be stopped early if the VE was greater than expected (**Figures 2, S3 Table S2**). If the true VE was 90%, nearly all of the trials would be stopped at the first interim analysis (1000 participants). If the true VE was 44.4%, approximately 25% of the trials would be stopped after 1000 participants were enrolled, 25% would stop with 2000 participants enrolled, and ∼25-30% would stop with 3000 participants enrolled. The incorporation of historical data into the analysis resulted in a small but consistent gain in power when the true VE was 44.4% (**Figures 2, S3**), despite the use of a more conservative threshold (**Table S2**). These numbers can be compared with a trial that used a single pre-determined sample size. With a single, final evaluation, 2400-2800 individuals would need to be enrolled in a new trial to achieve 80% power to detect a VE of 44.4%, depending on the prior distribution used.

Using a more conservative stopping rule (i.e., posterior probability of VE > 30%) resulted in a relatively small number of trials reaching the stopping criteria when there is a true, moderate effect of 44.4% (**Figure S4**). With a true, strong VE of 90%, almost all trials were still declared a success with just 1000 individuals enrolled.

We also evaluated trials where the threshold for stopping the trial was not adjusted to reduce the type I error rate. With this less stringent threshold for stopping at interim analyses (α_interim_=0.05), the models using historical information had a clear advantage in efficiency, with 72% of trials stopping for efficacy (P(VE > 0% | data) > 0.95) with just 1000 participants, compared with 43% of trials if data from previous studies were ignored (**Figure S5**).

## DISCUSSION

In this study, we considered two complementary approaches for conducting clinical trials more efficiently: (1) incorporating data from previous trials, and/or (2) the use of Bayesian group sequential trials. We found that if there is a need to maintain a low type I error rate, as in clinical trials used for licensure, incorporating data from previous trials yielded only a modest improvement in power. This is in line with previous findings [21]. If there is not a need to strictly control the type I error rate, as might be the case in a confirmatory trial or a post-licensure evaluation of vaccine effectiveness, incorporating historical data into the analysis would allow for more rapid assessments with smaller sample size. Group sequential designs with frequent interim analyses, with or without incorporating historical data, required smaller numbers of participants on average compared with trials with a single intervention, particularly when the intervention effect was strong.

This study builds upon well-developed methods for incorporating historical information into analyses of new data using dynamic borrowing prior distributions. There are a number of dynamic borrowing prior distributions that have been previously introduced, with the main categories including power priors [7,22], commensurate priors [6], and robust meta-analytic-predictive priors [23]. For our analyses, commensurate priors have the advantages of being easy to implement using Markov chain Monte Carlo sampling techniques while also being more flexible than the original power prior by allowing each parameter (e.g., intercept, slope) to borrow historical data at different rates as needed. We integrated data from a single historical trial with simulated data from a new hypothetical study. There are situations where it would be beneficial to integrate data from multiple historical estimates with new data. In this situation, more complex multi-source exchangeability models need to be used that determine whether to borrow information from one, more than one, or none of the historical trials [24].

Our results suggest that group sequential designs that incorporate historical data could have some advantages in terms of efficiency. However, there are some practical challenges to implementing the proposed methods in the context of a trial used for licensure. Regulators are often skeptical of the use of Bayesian methods for licensure trials [25]. The use of simulation studies, as we have done here, can help demonstrate the operating characteristics of the trial (e.g., power, type I error rate). The evaluation of the performance of a suite of prior distributions provides the most holistic, credible view of performance [26].

We compared analyses that used a stringent threshold to maintain a low type I error rate with analyses that used a less stringent threshold. There has been some debate about the need to account for type I error with Bayesian trials [27]. Some have argued that the Bayesian paradigm is not compatible with the concept of having a multitude of theoretical trials over which performance is evaluated. Similarly, with a Bayesian approach, it is natural to report and make decisions based on a continuous value of the posterior probability rather than a binary hypothesis test. While these arguments are valid, FDA guidance documents clearly state that Bayesian trials need to evaluate frequentist operating characteristics (power and type I error) [28]. If the thresholds used to declare ‘success’ of the trial are not adjusted, a large proportion of trials could declare success when there is no real effect when performing group sequential trials. Our analyses use a simple adjustment to reduce the type I error rate, using the same, more conservative threshold at each evaluation point [9]. More sophisticated adjustments that use more conservative thresholds at earlier time points could also be implemented [29], or a less stringent threshold could be used at the final evaluation point if the trial was not stopped at an interim analysis [2].

In these analyses, we focus on vaccines against RSV. While several vaccines are now licensed, there are many additional vaccine candidates in different stages of clinical development [30]. Therefore, issues around trial design will continue to be relevant for RSV vaccines in the coming years. When designing these trials, key questions will revolve around analytical methods, as we have discussed in this study, as well as questions around selection of primary clinical endpoint(s) and selection of study sites that will provide information that is useful for both regulators and country-level advisory committees that develop vaccine recommendations.

The general analytic methods evaluated here for clinical trials could have other applications, for example by incorporating data from the trials into post-licensure studies that monitor vaccine effectiveness and safety. In a post-licensure evaluation, the goal is typically estimation of effects rather than hypothesis testing [31]. Therefore, the need to use a more conservative threshold for significance is less important. There is also stronger prior information (from the phase 3 trials).

These types of approaches may be particularly valuable for quantifying vaccine effectiveness against rare but clinically important outcomes (e.g., severe disease or death) or evaluating potential safety signals. For instance, regulators have requested further monitoring of rates of preterm birth following the introduction of a new maternal vaccine against RSV [32]. Estimates from the phase 3 trial could be incorporated with observational studies to provide more rapid assessments of safety in a real-world setting. Further methodological work is needed to evaluate the performance of these methods with real-world data when there is also a need to adjust for potential confounders, and the estimand of interest is often an odds ratio rather than a risk ratio.

Our analyses have certain limitations. In real clinical trials, there would typically be stopping criteria based on efficacy, futility (evidence that the intervention is not effective), and safety. For simplicity, we focus on a stopping criterion based only on efficacy. We also use a simple criterion for stopping based on efficacy, with the same threshold at all interim analyses and on the final analysis. More sophisticated ‘spending rules’ that adjust α_interim_ over the course of the trial could lead to a more efficient trial but are unlikely to influence the conclusions of this study. Our analyses focused on maximizing trial efficiency (i.e., stopping as early as possible due to demonstrated efficacy). However, efficiency is not the only consideration when designing a trial. Ensuring the trial is sufficiently powered to detect safety signals is also important. Moreover, stopping earlier might make it more difficult to evaluate efficacy in particular subgroups or geographic regions, which might be important in the post-licensure context when advisory bodies are making recommendations for use.

In conclusion, group sequential analyses with frequent evaluations of efficacy, potentially with the incorporation of historical data, could allow for more efficient evaluations of vaccines and increase the chances that a trial is successful compared to a trial with a single pre-specified evaluation point. Given the high burden of disease caused by RSV and the recent track record of having a failed trial, these alternative designs should be considered for future evaluations of interventions against RSV as well as other endemic diseases.

## Data Availability

All of the code require to replicate the simulated data are available from: https://github.com/weinbergerlab/sequential_bayesian_trials

## Acknowledgements

Thanks to the extensive resources of Frank Harrell’s websites, which inspired some of the analyses here: https://hbiostat.org/bayes/; https://www.fharrell.com/post/bayes-seq

## Supplementary Figures

**Figure S1.**
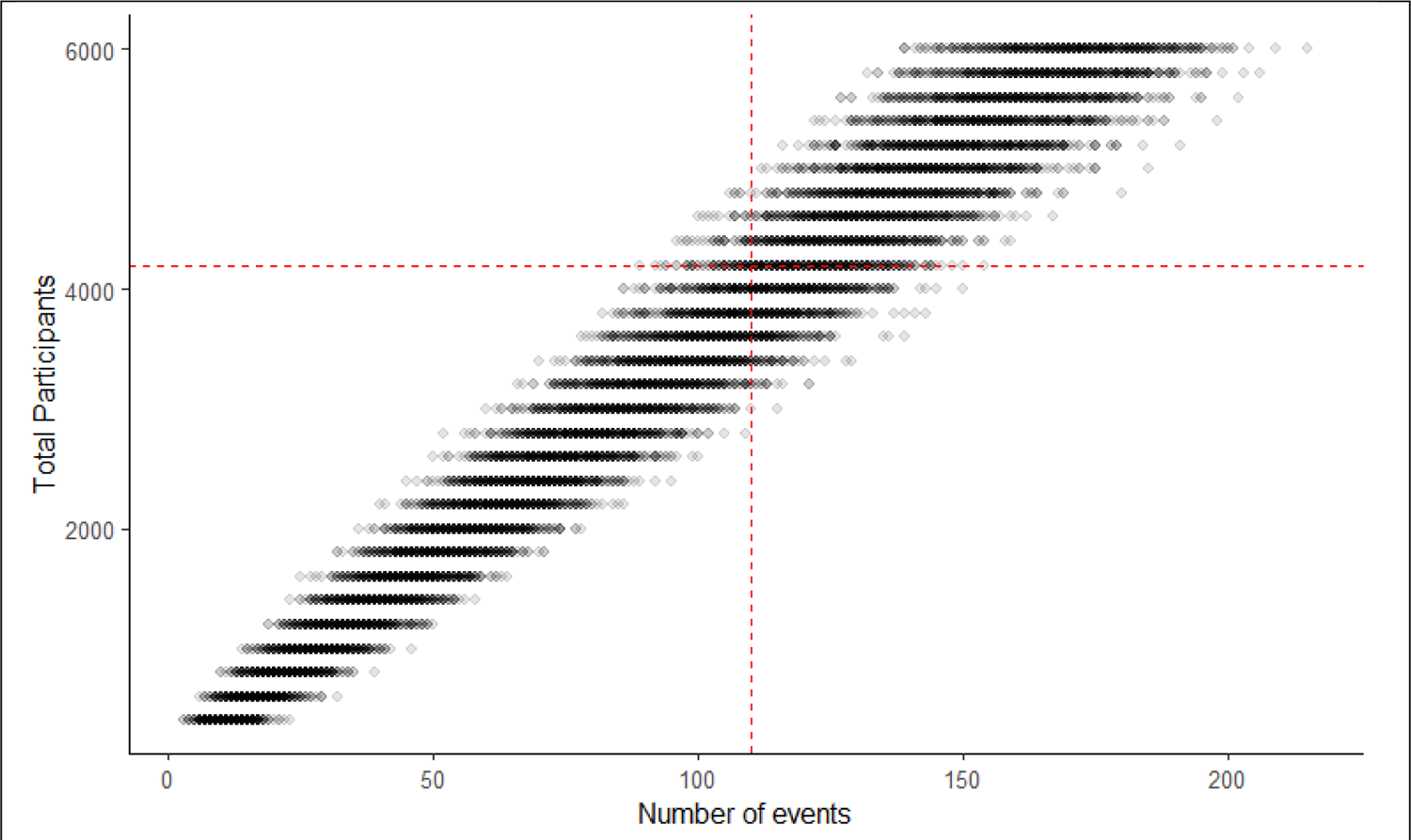
Relationship between average number of participants enrolled across both arms and the number of events in the trial. The red lines show the observed values used in the analysis from the Prepare trial.

**Figure S2.**
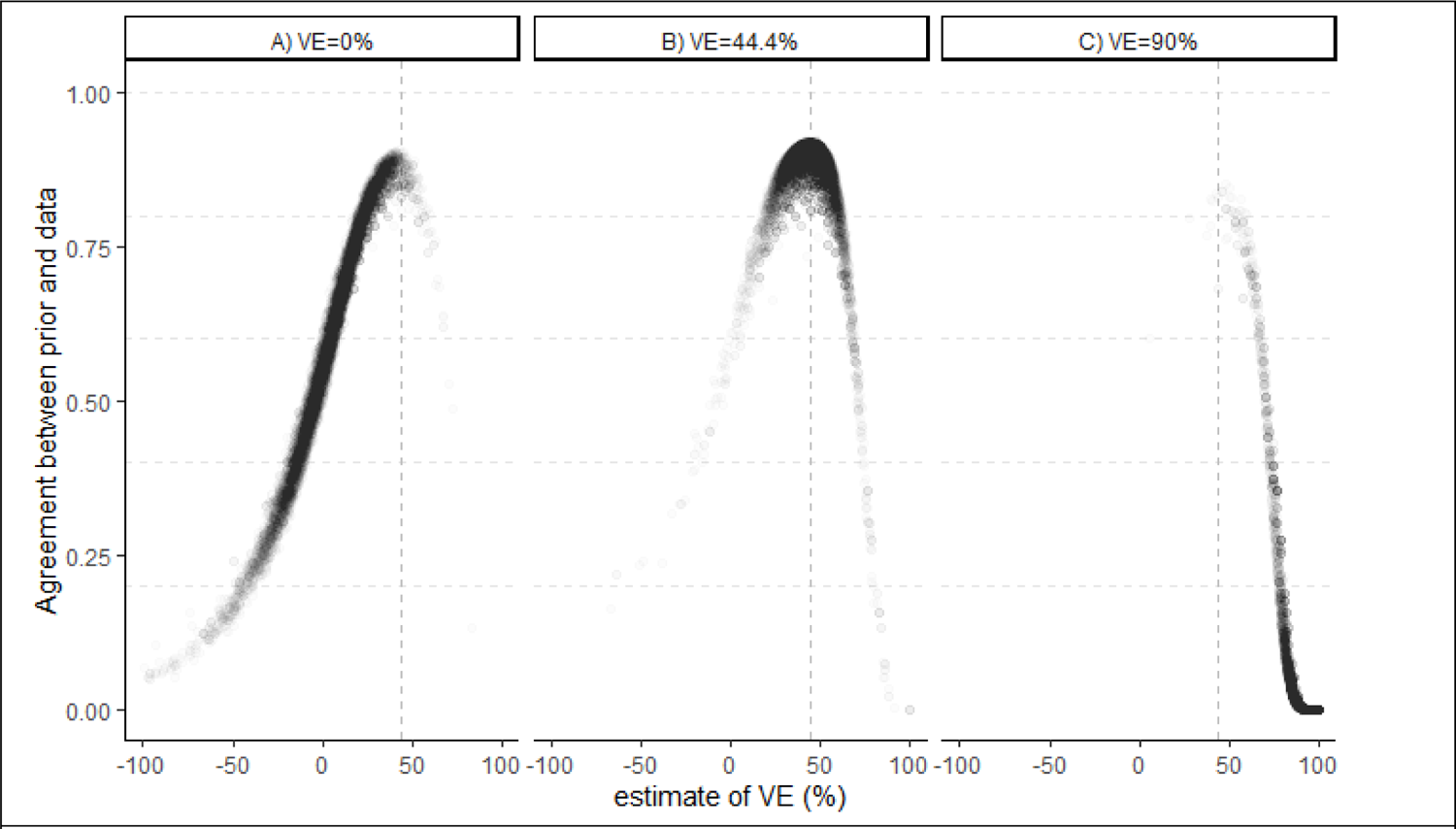
Relationship between estimated VE and the degree of data sharing between the original trial and the new trial. The values on the Y-axis represent a transformation of the variance parameter from the commensurate prior (exp(-σ^2^1)). If the variance is small, that indicates increased sharing and a transformed value close to 1. If the variance is large, this indicates decreased sharing, and the value approaches 0. The point estimate from the original trial (44.4%) is denoted with a dashed line and corresponds to where the agreement between the original and new trial data is greatest. Each dot is from a single simulated trial at a particular time point. Each panel corresponds to simulated data where different true VE levels (0%,44.4%, 90%) were used to generate the data.

**Figure S3.**
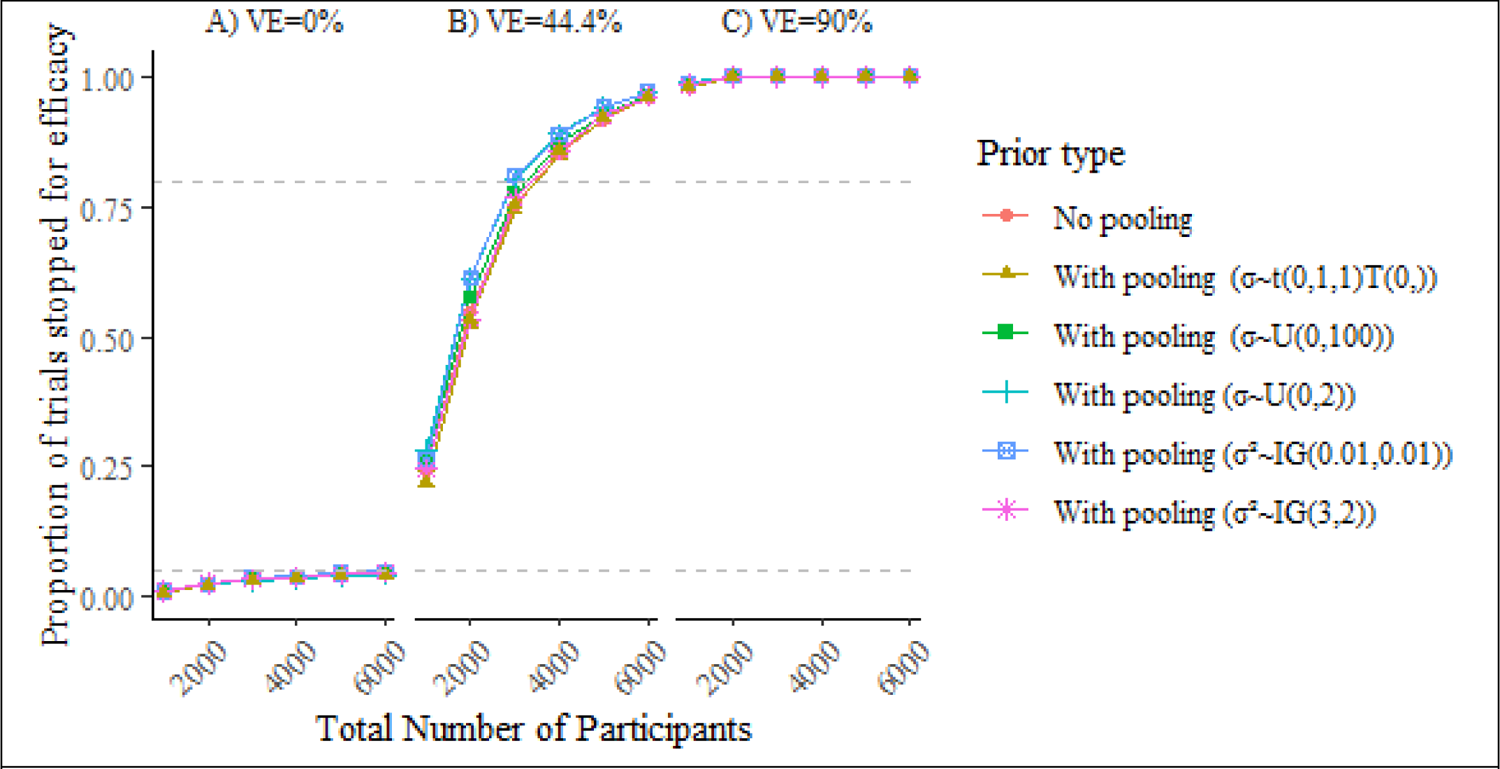
Cumulative proportion of trials that are declared a success (VE > 0%) using a threshold for stopping (α_interim_) that is adjusted to maintain a type I error rate of 5% for the study (α_total_). This shows the full set of models that were evaluated. VE is evaluated after every 1000 participants complete the follow-up period. From left to right, we consider scenarios with a “true” vaccine efficacy of (A) 0%, (B) 44.4%, and (C) 90%. The horizontal dashed lines are at 0.05 and 0.8. The horizontal dashed lines are at 0.05 and 0.80, which represent arbitrary thresholds for acceptable type I error rates and power, respectively. The value of α_interim_ is adjusted so that the proportion of trials stopped for efficacy is 0.05 at n=6000 participants when the true VE=0% (left panel).

**Figure S4.**
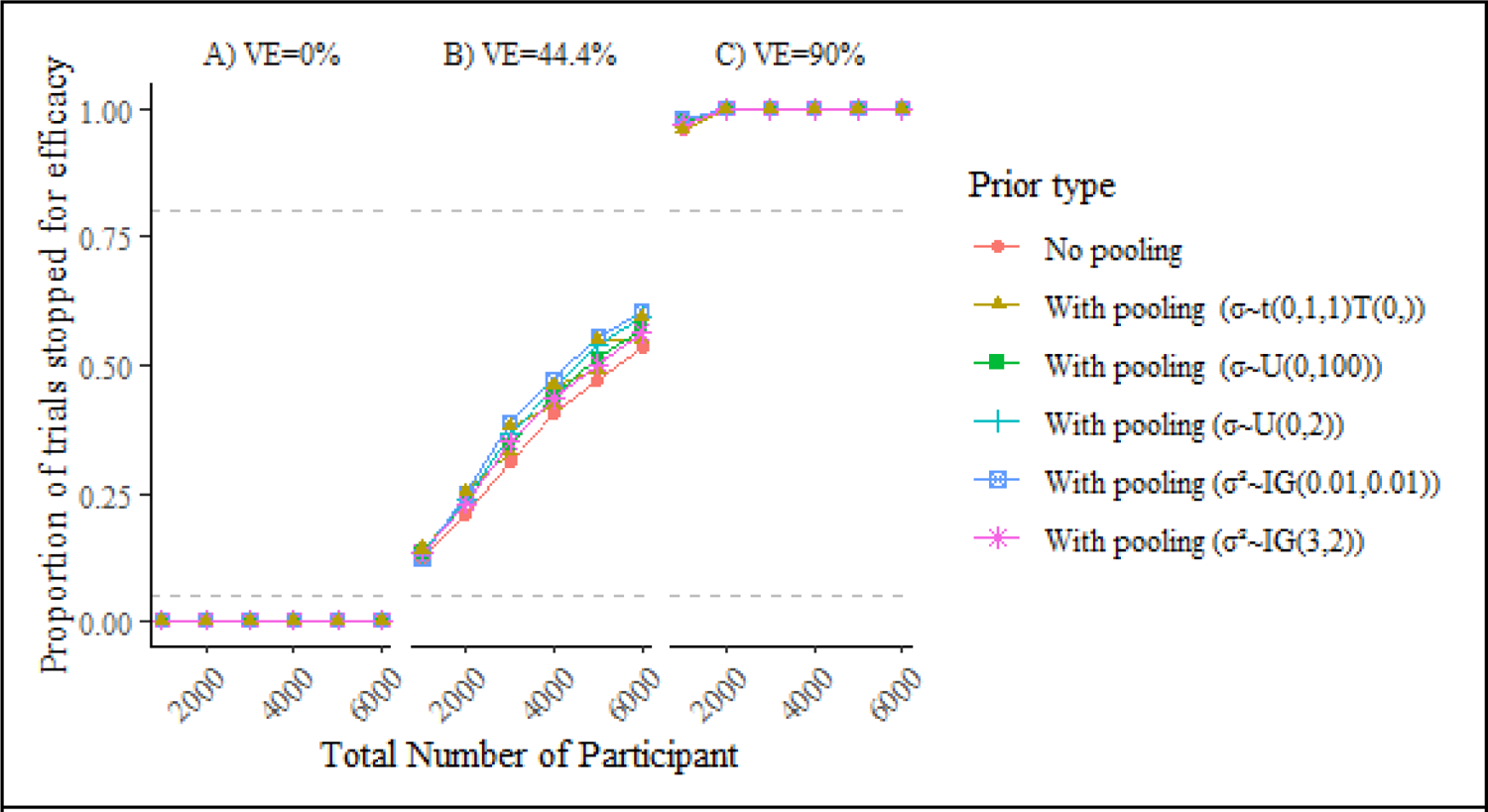
Cumulative proportion of trials that are declared a success, using a more stringent definition of success (VE > 30%) and a threshold for stopping (α_interim_) that is set to 0.05. (A) VE evaluated after every 1000 participants complete the follow up period. From left to right, we consider scenarios with a “true” vaccine efficacy of (A) 0%, (B) 44.4%, and (C) 90%. The horizontal dashed lines are at 0.05 and 0.80, which represent arbitrary thresholds for acceptable type I error rates and power, respectively. Because the crierion for success was more stringent, it was not necessary to adjust α_interim_ to a smaller value to maintain an overall α_total_ of 0.05.

**Figure S5.**
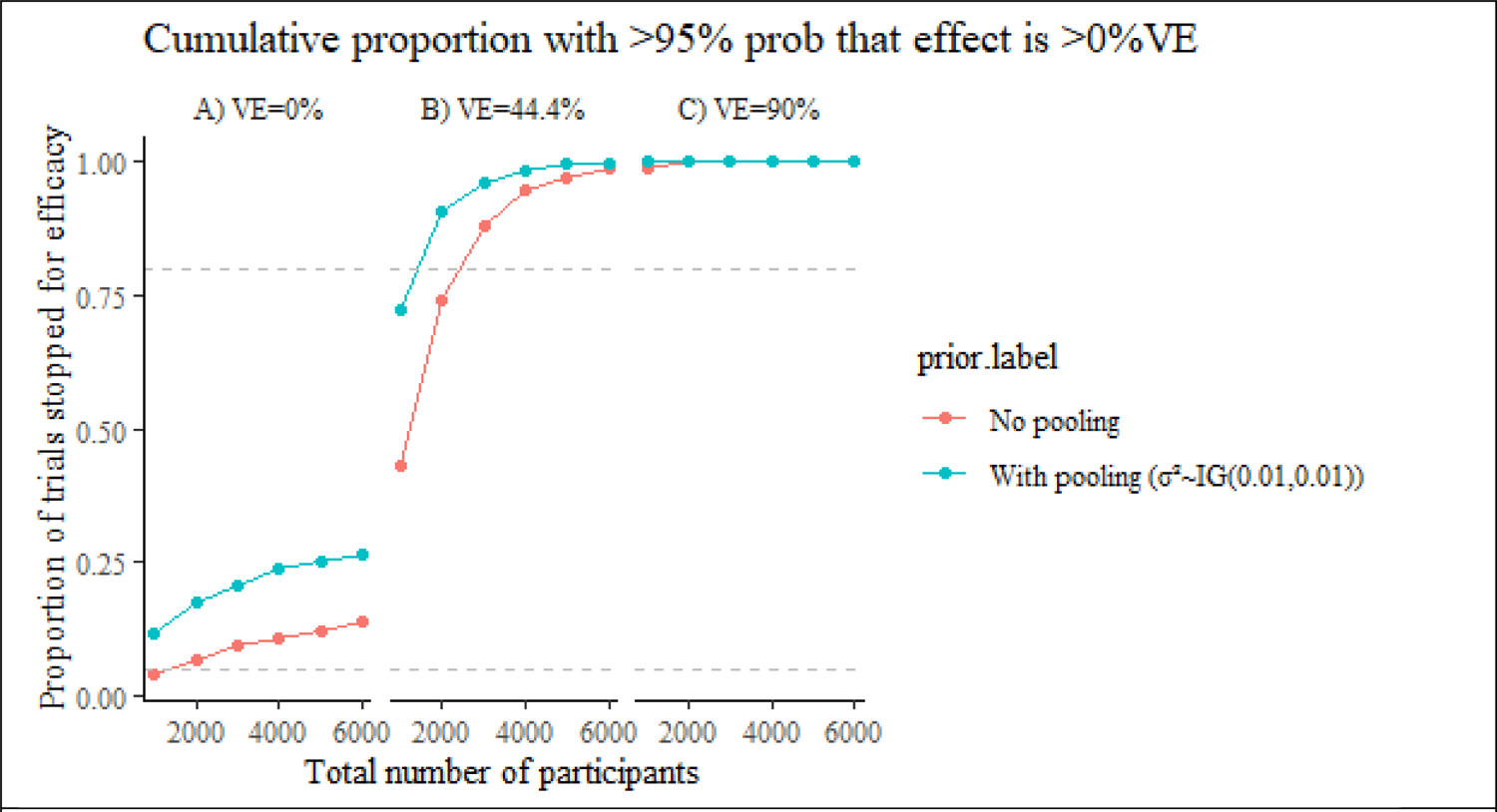
Cumulative proportion of trials that are declared a success using a less stringent threshold for stopping (α_interim_=0.05). Unlike other analyses shown in the manuscript, there is no adjustment of α_interim_ to reduce type I error (i.e. P(VE > 0% | data) > 0.95). VE is evaluated after every 1000 participants complete the follow-up period. From left to right, we consider scenarios with a “true” vaccine efficacy of (A) 0%, (B) 44.4%, and (C) 90%. Panel A shows that up to 25% of trials would be stopped early and (falsely) conclude the vaccine is efficacious even when there is no effect in the new trial. The horizontal dashed lines are at 0.05 and 0.80, which represent arbitrary thresholds for acceptable type I error rates and power, respectively.

---

**Table S1.** Summary of priors used in the analyses.

| Prior type | Prior on vaccine effect ($\beta_1$)* |
| --- | --- |
| No pooling (Minimally informative prior) | $N(0, 100^2)$ |
| No pooling (Skeptical prior) | $N(0, 3.32)$ |
| Pooling (Commensurate prior) | $N(\delta_1, \sigma^2); \sigma_1 \sim \text{Unif}(0, 2)$ |
| Pooling (Commensurate prior) | $N(\delta_1, \sigma^2); \sigma^2_1 \sim \text{IG}(0.01, 0.01)$ |

**Table S2.**
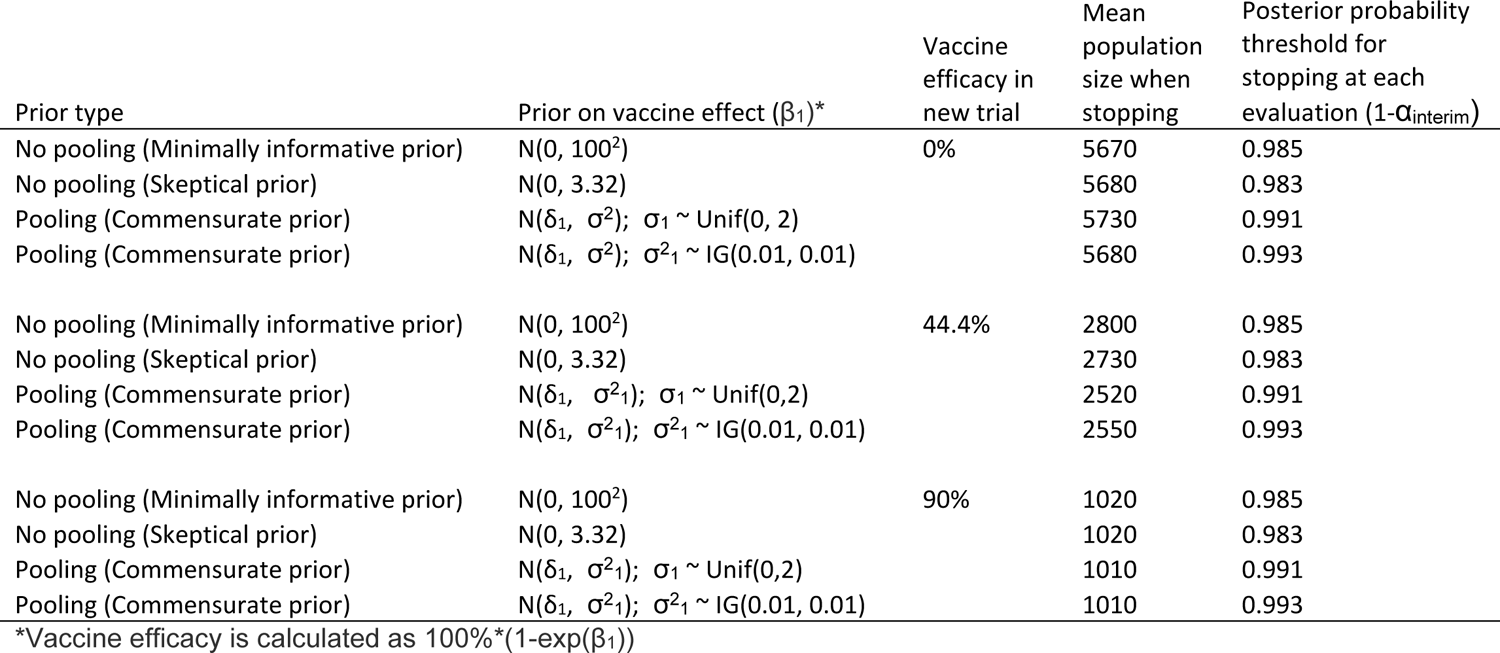
Average final population size when using a stopping criterion that maintains a type I error of 5% for the trial, evaluating P(VE > 0|data).

